# DIFFERENTIAL IMPROVEMENTS BETWEEN MEN AND WOMEN IN REPEATED CROSSFIT® OPEN WORKOUTS

**DOI:** 10.1101/2023.03.22.23287575

**Authors:** Gerald T. Mangine, Nina Grundlingh, Yuri Feito

## Abstract

The CrossFit® Open (CFO) acts a preliminary round that qualifies men and women for later stages of its annual Games competition. The CFO typically consists of 4-6 workouts that variably challenge an athlete’s weightlifting strength, gymnastic skill, and endurance capacity. Except for differences in prescribed intensity loads, workouts are designed the same for men and women to elicit a similar challenge. While all workouts within a single year are unique to each other, one has been repeated from a previous CFO each year between 2012 and 2021. Because previous CFO workouts are often integrated into training, improvements are expected when a workout is officially repeated. However, besides documented record performances, it is unclear whether most athletes are improving, if these improvements affect ranking, or if differences exist between men and women.

**PURPOSE:** To examine sex differences and performance changes across repeated CFO workouts, as well as their effect on CFO and workout ranking.

**METHODS:** Eleven separate samples of 500 men and 500 women, who were representative of the same overall percent rank within each year involving one of the nine repeated CFO workouts (2011-2021) were drawn for this study. Each athlete’s age (18-54 years), rank (overall and within each workout), and reported workout scores were collected from the competition’s publicly-available leaderboard. Each sample had excluded any athlete who had not met minimum performance criteria (e.g., at least one completed round) for all prescribed (Rx) workouts within a given year (including those not analyzed). Since some workouts could be scored as repetitions completed or time-to-completion (TTC), and because programming was often scaled between men and women, all scores were converted to a repetition completion rate (repetitions divided by TTC [in minutes]).

**RESULTS:** Separate sex x time analyses of variance with repeated measures revealed significant (*p* < 0.05) interactions in all but one (CFO 18.4 vs. 20.3) repeated workout comparison. Initially, men were faster in four workouts (∼18.5%, range = 3.9 – 35.0%, *p* < 0.001), women in two (∼7.1%, range = 5.2 – 9.0%, *p* < 0.001), and they tied in the remaining three workouts. When these workouts were repeated in subsequent years, men were no longer faster in two workouts (CFO 11.1 to 14.1 and CFO 12.4 to 13.3) but became faster in another (CFO 16.4 to 17.4). In contrast, women were slower in CFO 14.2 and became faster than men when the workout repeated (CFO 15.2), but then performed CFO 19.2 slower than men, a workout they initially completed faster (CFO 16.2). Though performance improved in seven of the nine workouts (∼14.3%, *p* < 0.001) and percentile rank was controlled, athletes earned a lower rank (overall and within workout) on each repeated workout (*p* < 0.001).

**CONCLUSIONS:** Performance (measured as repetition completion rate) has improved in most repeated CFO workouts, particularly females. However, improvements seen among all athletes, along with increased participation, have made it more difficult for athletes to improve their overall rank. To rank higher, individual athlete must improve their pace to a greater degree than the average improvements seen across the competitive field.

## INTRODUCTION

Developed in 2000, CrossFit® is a training strategy that incorporates a variety of functional, multimodal movements into workouts meant to be performed at a relatively high-intensity for the purpose of improving general physical preparedness (1-3). The strategy aims to avoid forming any linear structure to its daily, weekly, and monthly programming, and instead, constantly varies the stimulus to promote simultaneous and generalized improvements in fitness (2). It became a sport in 2007 when the first CrossFit® Games competition was held on a ranch in Aromas, California (3). Though it and the following year’s competitions were open to anyone who could travel to the ranch, the sport quickly grew in popularity, which necessitated the introduction of preliminary, qualifying rounds. In 2011, the sport had grown so large that an online, qualification round known as the CrossFit® Open (CFO) was introduced (3). It was needed to reduce the initial participant pool of 26,000 to the top 60 athletes within 17 worldwide regions, with the top 1 – 3 athletes from each region progressing to the Games. While several changes to the competition’s structure have been made since 2011 (3), the existence of the CFO has remained. The 3 – 5-week CFO has typically consisted of 4 – 6 workouts that variably challenge an athlete’s weightlifting strength, gymnastic skill, and endurance capacity (4, 5). The details of each workout are announced weekly, and competitors may rely on the fact that all workouts within a particular year will be unique to each other. However, except for in 2011 and 2022, each CFO competition has included a workout drawn from a previous competition, with nine distinct workouts acting as repeats between 2012 and 2021. Adapted from (4), the composition and movement standards for these workouts are presented in Tables 1 and 2, respectively.

**Table 1.** Repeated CrossFit® Open Workouts.

| Duration | Details | Appearances (scoring) |  |  |
| --- | --- | --- | --- | --- |
| 10-minute AMRAP | 30 x Double-unders<br>15 x Power snatches (75lbs / 55lbs) | 11.1<br>(repetitions) | 14.1<br>(repetitions) |  |
| 7-minute AMRAP | 3 x Thrusters (100lbs / 65lbs)<br>3 x Chest-to-bar pull-ups<br>*\*Add 3 repetitions after each completed round* | 11.6<br>(repetitions) | 12.5<br>(repetitions) | 18.5<br>(repetitions) |
| 12-minute AMRAP | 150 x Wall ball shots (20lbs / 14lbs to 10' / 9' target)<br>90 x Double-unders<br>30 x Muscle-ups | 12.4<br>(repetitions) | 13.3<br>(repetitions) |  |
| 3-minute rounds<br>(indefinite) | *Complete two sets of:*<br>10 x Overhead squats (95lbs / 65lbs)<br>10 x Chest-to-bar pull-ups<br>*\*Add 3 minutes and 2 repetitions for each completed round* | 14.2<br>(repetitions) | 15.2<br>(repetitions) |  |
| TTC | *21-18-15-12-9-6-3 repetitions of:*<br>Thrusters (95lbs / 65lbs)<br>Bar-facing burpees | 14.5 (TTC) | 16.5 (TTC) |  |
| 4-minute rounds (20-minute time limit) | 25 x Toes-to-bar<br>50 x Double-unders<br>Squat cleans (R1: 135lbs / 85lbs x 15; R2: 185lbs / 115lbs x 13; R3: 225lbs / 145lbs x 11; R4: 275lbs / 175lbs x 9; R5: 315lbs / 205lbs x 7)<br>*\*Add 4 minutes for each completed round* | 16.2 (TTC or repetitions) | 19.2 (TTC or repetitions) |  |
| 13-minute AMRAP | 55 x Deadlifts (225lbs / 155lbs)<br>55 x Wall ball shots (20lbs / 14lbs to 10' / 9')<br>55 x Calorie rowing<br>55 x Handstand push-ups | 16.4<br>(repetitions) | 17.4<br>(repetitions) |  |
| 20-minute time limit | *Alternate the following:*<br>Dumbbell snatches (50lbs / 35lbs) x 10-20-30-40-50 repetitions<br>15 x Burpee box jump-overs (24" / 20") | 17.1 (TTC or repetitions) | 21.2 (TTC or repetitions) |  |
| 9-minute time limit | *Complete 21-15-9 repetitions of:*<br>Deadlifts (225lbs / 155lbs)<br>Handstand push-ups<br>*Then complete 21-15-9 repetitions of:*<br>Deadlifts (315lbs / 205lbs)<br>Handstand walk (50') | 18.4 (TTC or repetitions) | 20.3 (TTC or repetitions) |  |

Notes: AMRAP = 'as many repetitions as possible'; TTC = time to completion

**Table 2.**
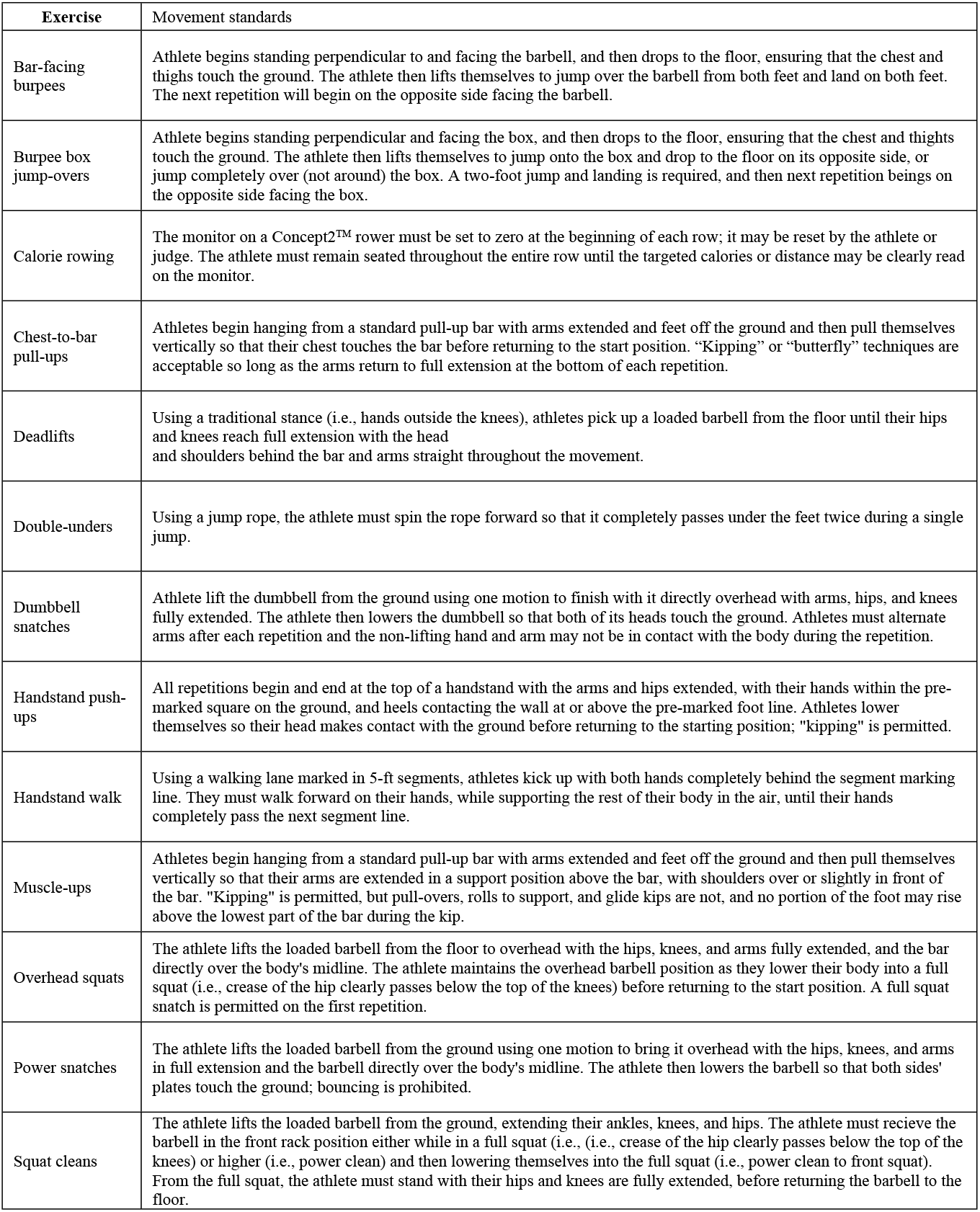
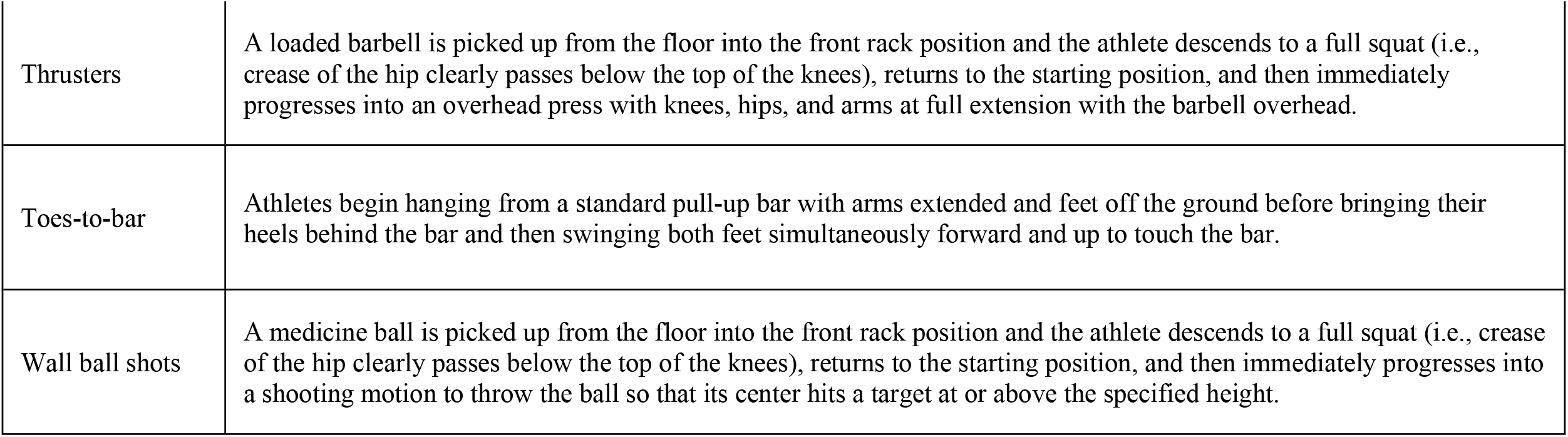
Exercise movement standards.

| Exercise | Movement standards |
| --- | --- |
| Bar-facing burpees | Athlete begins standing perpendicular to and facing the barbell, and then drops to the floor, ensuring that the chest and thighs touch the ground. The athlete then lifts themselves to jump over the barbell from both feet and land on both feet. The next repetition will begin on the opposite side facing the barbell. |
| Burpee box jump-overs | Athlete begins standing perpendicular and facing the box, and then drops to the floor, ensuring that the chest and thighs touch the ground. The athlete then lifts themselves to jump onto the box and drop to the floor on its opposite side, or jump completely over (not around) the box. A two-foot jump and landing is required, and then next repetition begins on the opposite side facing the box. |
| Calorie rowing | The monitor on a Concept2™ rower must be set to zero at the beginning of each row; it may be reset by the athlete or judge. The athlete must remain seated throughout the entire row until the targeted calories or distance may be clearly read on the monitor. |
| Chest-to-bar pull-ups | Athletes begin hanging from a standard pull-up bar with arms extended and feet off the ground and then pull themselves vertically so that their chest touches the bar before returning to the start position. "Kipping" or "butterfly" techniques are acceptable so long as the arms return to full extension at the bottom of each repetition. |
| Deadlifts | Using a traditional stance (i.e., hands outside the knees), athletes pick up a loaded barbell from the floor until their hips and knees reach full extension with the head and shoulders behind the bar and arms straight throughout the movement. |
| Double-unders | Using a jump rope, the athlete must spin the rope forward so that it completely passes under the feet twice during a single jump. |
| Dumbbell snatches | Athlete lift the dumbbell from the ground using one motion to finish with it directly overhead with arms, hips, and knees fully extended. The athlete then lowers the dumbbell so that both of its heads touch the ground. Athletes must alternate arms after each repetition and the non-lifting hand and arm may not be in contact with the body during the repetition. |
| Handstand push-ups | All repetitions begin and end at the top of a handstand with the arms and hips extended, with their hands within the pre-marked square on the ground, and heels contacting the wall at or above the pre-marked foot line. Athletes lower themselves so their head makes contact with the ground before returning to the starting position; "kipping" is permitted. |
| Handstand walk | Using a walking lane marked in 5-ft segments, athletes kick up with both hands completely behind the segment marking line. They must walk forward on their hands, while supporting the rest of their body in the air, until their hands completely pass the next segment line. |
| Muscle-ups | Athletes begin hanging from a standard pull-up bar with arms extended and feet off the ground and then pull themselves vertically so that their arms are extended in a support position above the bar, with shoulders over or slightly in front of the bar. "Kipping" is permitted, but pull-overs, rolls to support, and glide kips are not, and no portion of the foot may rise above the lowest part of the bar during the kip. |
| Overhead squats | The athlete lifts the loaded barbell from the floor to overhead with the hips, knees, and arms fully extended, and the bar directly over the body's midline. The athlete maintains the overhead barbell position as they lower their body into a full squat (i.e., crease of the hip clearly passes below the top of the knees) before returning to the start position. A full squat snatch is permitted on the first repetition. |
| Power snatches | The athlete lifts the loaded barbell from the ground using one motion to bring it overhead with the hips, knees, and arms in full extension and the barbell directly over the body's midline. The athlete then lowers the barbell so that both sides' plates touch the ground; bouncing is prohibited. |
| Squat cleans | The athlete lifts the loaded barbell from the ground, extending their ankles, knees, and hips. The athlete must receive the barbell in the front rack position either while in a full squat (i.e., crease of the hip clearly passes below the top of the knees) or higher (i.e., power clean) and then lowering themselves into the full squat (i.e., power clean to front squat). From the full squat, the athlete must stand with their hips and knees are fully extended, before returning the barbell to the floor. |
| Sex differences in repeated CFO workouts 27 |  |
| Thrusters | A loaded barbell is picked up from the floor into the front rack position and the athlete descends to a full squat (i.e., crease of the hip clearly passes below the top of the knees), returns to the starting position, and then immediately progresses into an overhead press with knees, hips, and arms at full extension with the barbell overhead. |
| Toes-to-bar | Athletes begin hanging from a standard pull-up bar with arms extended and feet off the ground before bringing their heels behind the bar and then swinging both feet simultaneously forward and up to touch the bar. |
| Wall ball shots | A medicine ball is picked up from the floor into the front rack position and the athlete descends to a full squat (i.e., crease of the hip clearly passes below the top of the knees), returns to the starting position, and then immediately progresses into a shooting motion to throw the ball so that its center hits a target at or above the specified height. |

Competitive success in CrossFit® is anecdotally thought to be dependent on an athlete’s proficiency in each of the numerous potential exercises that may appear in workouts, and all associated physiological traits. This idea is supported by several studies reporting relationships between performance and most investigated physiological variables (6-12). While the collective findings of these studies may ultimately prove useful for developing generalized training recommendations, they all suffer from the same underlying assumption that the needs of CrossFit® athletes are static. Alongside and within some of these studies, it has also been suggested that the relevance of these traits may be modulated by the athlete’s experience (7, 12-14). As one becomes more familiar with the physical tasks appearing in sport, they learn to activate more muscle when given the same tasks and eliminate inefficient and unnecessary actions (15-17). An experienced athlete would have had more opportunities to develop relevant skills and strategies that affect their approach to a workout (i.e., pacing strategy) and its resultant physiological demand. That is, the more experienced athlete might complete more work within a given duration because of a more effective pacing strategy. This is relevant to CrossFit® competition because workouts are commonly structured to produce a score that emphasizes maximal workout density (i.e., complete assigned work as fast as possible or complete as much work as possible within the assigned time) (1, 2). Athletes are routinely scored by either their time to complete (TTC) assigned work or their ability to complete ‘as many repetitions as possible’ (AMRAP) of assigned work within a specified time limit. After their introduction, CFO workouts become benchmarks (i.e., named workouts that are commonly known across training facilities) that may be integrated into training. It is reasonable to expect that greater familiarity with these workouts would lead to improved scores whenever one is officially repeated in competition (18, 19). However, aside from documented record performances from the sport’s top athletes (4, 5), it is unclear whether this is true for the remaining competition pool. Further, the impact of increased participation on performance and ranking is also unknown and a worthy consideration.

The growth in female participation is another important consideration. The initial CFO pool of 26,000 athletes consisted of 4,506 women (34.3% of all athletes) and that number grew to 42,799 women in 2021 (36.7% of non-scaled or age-group athletes who completed all CFO workouts as prescribed) (20). Unlike more traditional sports, gender equity is purposely emphasized in CrossFit®. The same number of men and women compete together on teams, the same number are invited to the Games, the same fans are spectators at both competitions, and male and female winners receive the same amount of monetary compensation (21). The only observable difference between the male and female competitions are participation and programming. With programming, workouts are often scaled between sexes to account for known physiological differences and deliver a similar challenge (4, 5), though scaling practices do not appear to be uniformly prescribed and their effect on performance has not been specifically explored. Among repeated CFO workouts, weightlifting intensity loads were the most often scaled programming feature with female loads being prescribed at 62.2 – 73.3% of the loads prescribed to men, along with wall ball shot target distance (3.05 m versus 2.74 m) and box jump height (61 cm versus 51 cm) (4). While scaling resistance-based exercises may be assumed to account for known strength and power differences between men and women (22), those differences were not considered (i.e., scaled) with gymnastic-calisthenic exercise prescription (e.g., handstand push-ups/walking, burpees, pull-ups, and muscle-ups). It might be assumed that scaling was considered unnecessary for these movements because they primarily require the athlete to maneuver their own body mass about an object (e.g., pull-up bar, rings, box) (1, 2) and women tend to possess less body mass (22, 23); thus, equating the difficulty. Scaling was also not applied to jumping rope and rowing tasks, which seemingly does not account for known sex differences in physiological measures of cardiorespiratory fitness (22, 24). It is unknown whether differences in programming have affected male and female CFO performance or if this has changed over time.

In non-CFO workouts, one study reported no differences between 13 men and 10 women for a 20-minute AMRAP that scaled rowing calories (men = 13, women = 11), deadlift loads (men = 62 kg, women = 44 kg), and kettlebell swing loads (men = 24 kg, women = 16 kg) but not burpees (25),. Likewise, no differences were found in a TTC version of the same workout, where participants completed sufficient rounds to equate the total volume of the 20-minute AMRAP. However, experience (the only available proxy of athlete skill) was significantly different between sexes, which could have affected how each approached the workouts.

Furthermore, the extremely small sample size limits the generalizability of these results to the overall CrossFit® population. More recently, normative scores were developed from very large random samples of CFO competitors (*n* = 7,046 – 89,792) for all workouts programmed between 2011 and 2022 (26), and sex differences were observed in 56 out of 60 total workouts. Although this implies CFO scaling between sexes has been ineffective (i.e., if scaling was properly applied, no differences should have been observed), any definitive conclusions would be premature at this time. The average sample for women was approximately 43.1% the size of the men’s samples, and it cannot be assumed that participant skill was equated between each sex’s sample on any given workout. Making comparisons between samples of equal size and equated skill would provide a better understanding about whether scaling practices have been effective (or in need of change). In turn, this information would assist in the development of generalized training recommendations and provide better context for studies aiming to determine physiological predictors of performance. Thus, the purpose of this investigation was to begin that process by examining sex differences and performance changes across repeated CFO workouts in similarly-ranked (i.e., equated skill) athletes. A secondary aim of this study was to determine how changes in performance translated to ranking in each workout in each workout and the overall competition. Based on pilot work (26, 27), we hypothesized that men would perform better on more workouts, regardless of scaling, but both sexes would improve equally over time. Nevertheless, improved performance would not translate to a higher official CFO competition rank due to increased overall participation.

## METHODS

### Study design

To determine sex differences and performance changes across repeated CFO workouts, performance data was collected for all athletes participating in CFO competitions from 2011 to 2021. All competition results were obtained from the JSON file located on the publicly-available, official competition leaderboard (20). Since these data were pre-existing and publicly available, the Kennesaw State University’s Institutional Review Board classified this study as exempt, and participants did not have to provide their informed consent. Python3 was used to convert the data into a CSV format and treated in Microsoft Excel (v. 365, Microsoft Corporation, Redmond, VA, USA). Treating the data involved removing all age-group athletes (e.g., teens and masters) and cases that did not meet study inclusion criteria. The retained data included each athlete’s age and final overall ranking (i.e., official CFO rank awarded within a given year), as well as their official rank and score for each repeated CFO workout that they completed. Subsequently, differences were examined between sexes and repeated efforts.

### Participants

Based on pilot data (27) and the expectation of a small effect (Effect of *f* = 0.10), a priori analysis using G*Power (v. 3.1.9.7, Heinrich-Heine-Universität, Germany) for a repeated-measures design indicated at least 328 participants would provide sufficient power (α = 0.05, β = 0.95) to observe differences between sexes and performances in each repeated CFO workout.

Since collecting data on the same 328 men and women from 2011 to 2021 would produce a very specific sample, one that is likely representative of a subset of CFO participants, this study identified cases based on percent rank and made comparisons between athletes of the same percent rank across competition years.

Initially, a stratified list of 500 percentile ranks were identified based on the approximate percentage of cases that would fall within each standard deviation (SD) bin: 0.0 – 0.5 SD (38.2%, *n* = 192), 0.5 – 1.0 SD (30.0%, *n* = 150), 1.0 – 1.5 SD (18.4%, *n* = 92), 1.5 – 2.0 SD (8.8%, *n* = 44), 2.0 – 2.5 SD (3.4%, *n* = 16), 2.5 – 3.0 SD (1.0%, n = 4), and 3.0 – 3.5 SD (0.2%, *n* = 2) (28). Ranks were evenly divided within each bin and across each side of the mean (e.g., – 2.75 SD, –2.5 SD, +2.5 SD, and +2.75 SD). These percent ranks would be used to identify the specific cases within each CFO year that would be drawn from the pool of athletes who also met study criteria. Age, rank, and performance data were retained for all athletes, between the ages of 18 and 54 years (i.e., non-age group athletes), who completed all CFO workouts as prescribed (i.e., as Rx with no within-sex scaling) within a specific competition year. Additionally, cases were excluded if they did not complete at least one round (in AMRAP-style workouts), the first exercise couplet (in repeated couplet workouts), or all repetitions assigned for the first exercise (for TTC or when multiple rounds were not expected) for every workout within a single competition year. These criteria were meant to limit the inclusion of workout “specialists” and those who did not intend on completing or could not perform the exercises for the Rx workout. To match specific cases with the identified percent ranks to be drawn from within each year’s athlete pool, retained athletes were ordered based on their final within-sex overall competition rank and then assigned a within-year percent rank (i.e., percent rank among athletes meeting study criteria). The final within-year percent ranking was used to identify 500 athletes within each year who would be included in this study. Thus, the same array of percent ranks was represented by the athletes retained on each year. Table 3 provides a summary of the initial pool of competitors for each year, the number of cases meeting study criteria, and the age and final competition ranking characteristics of those retained for analysis.

**Table 3.**
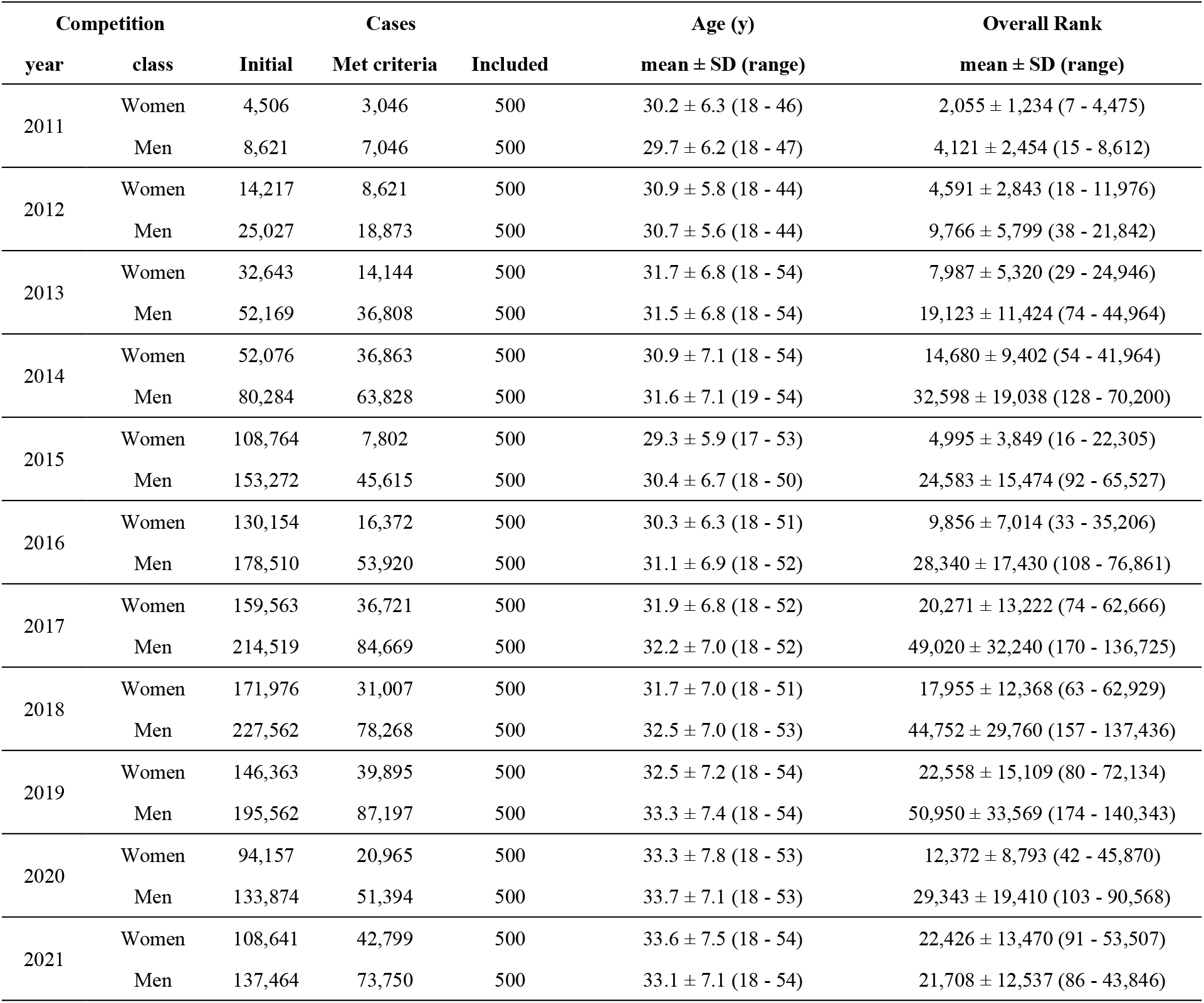
Case selection and sample characteristics.

| Competition |  | Cases |  |  | Age (y) | Overall Rank |
| --- | --- | --- | --- | --- | --- | --- |
| year | class | Initial | Met criteria | Included | mean ± SD (range) | mean ± SD (range) |
| 2011 | Women | 4,506 | 3,046 | 500 | 30.2 ± 6.3 (18 - 46) | 2,055 ± 1,234 (7 - 4,475) |
| | Men | 8,621 | 7,046 | 500 | 29.7 ± 6.2 (18 - 47) | 4,121 ± 2,454 (15 - 8,612) |
| 2012 | Women | 14,217 | 8,621 | 500 | 30.9 ± 5.8 (18 - 44) | 4,591 ± 2,843 (18 - 11,976) |
| | Men | 25,027 | 18,873 | 500 | 30.7 ± 5.6 (18 - 44) | 9,766 ± 5,799 (38 - 21,842) |
| 2013 | Women | 32,643 | 14,144 | 500 | 31.7 ± 6.8 (18 - 54) | 7,987 ± 5,320 (29 - 24,946) |
| | Men | 52,169 | 36,808 | 500 | 31.5 ± 6.8 (18 - 54) | 19,123 ± 11,424 (74 - 44,964) |
| 2014 | Women | 52,076 | 36,863 | 500 | 30.9 ± 7.1 (18 - 54) | 14,680 ± 9,402 (54 - 41,964) |
| | Men | 80,284 | 63,828 | 500 | 31.6 ± 7.1 (19 - 54) | 32,598 ± 19,038 (128 - 70,200) |
| 2015 | Women | 108,764 | 7,802 | 500 | 29.3 ± 5.9 (17 - 53) | 4,995 ± 3,849 (16 - 22,305) |
| | Men | 153,272 | 45,615 | 500 | 30.4 ± 6.7 (18 - 50) | 24,583 ± 15,474 (92 - 65,527) |
| 2016 | Women | 130,154 | 16,372 | 500 | 30.3 ± 6.3 (18 - 51) | 9,856 ± 7,014 (33 - 35,206) |
| | Men | 178,510 | 53,920 | 500 | 31.1 ± 6.9 (18 - 52) | 28,340 ± 17,430 (108 - 76,861) |
| 2017 | Women | 159,563 | 36,721 | 500 | 31.9 ± 6.8 (18 - 52) | 20,271 ± 13,222 (74 - 62,666) |
| | Men | 214,519 | 84,669 | 500 | 32.2 ± 7.0 (18 - 52) | 49,020 ± 32,240 (170 - 136,725) |
| 2018 | Women | 171,976 | 31,007 | 500 | 31.7 ± 7.0 (18 - 51) | 17,955 ± 12,368 (63 - 62,929) |
| | Men | 227,562 | 78,268 | 500 | 32.5 ± 7.0 (18 - 53) | 44,752 ± 29,760 (157 - 137,436) |
| 2019 | Women | 146,363 | 39,895 | 500 | 32.5 ± 7.2 (18 - 54) | 22,558 ± 15,109 (80 - 72,134) |
| | Men | 195,562 | 87,197 | 500 | 33.3 ± 7.4 (18 - 54) | 50,950 ± 33,569 (174 - 140,343) |
| 2020 | Women | 94,157 | 20,965 | 500 | 33.3 ± 7.8 (18 - 53) | 12,372 ± 8,793 (42 - 45,870) |
| | Men | 133,874 | 51,394 | 500 | 33.7 ± 7.1 (18 - 53) | 29,343 ± 19,410 (103 - 90,568) |
| 2021 | Women | 108,641 | 42,799 | 500 | 33.6 ± 7.5 (18 - 54) | 22,426 ± 13,470 (91 - 53,507) |
| | Men | 137,464 | 73,750 | 500 | 33.1 ± 7.1 (18 - 54) | 21,708 ± 12,537 (86 - 43,846) |

### Workout descriptions

The specific programming details for each workout included in this study are provided in Tables 1 and 2. During CFO competition, workouts were individually released on each week of the competition, and athletes have predominantly been given four days to submit their best score to competition officials (e.g., Thursday evening to Monday evening). To be recognized as valid, any submitted attempt must have either been completed at a CrossFit® affiliate in front of a judge who passed the online Judge’s Course, or filmed using standardized criteria (5). Once a submission period ends, competition officials review and certify attempts, and award each a final rank and points. Because all data was collected from the official competition leaderboard (20), this study assumed that all workout criteria and movement standards had been met and verified by competition officials. The data retained for analysis included the athlete’s official rank for each workout and score, recorded as TTC or repetitions. For uniformity and to enable fair comparisons when a workout’s official score could be stated as TTC or repetitions completed (i.e., when a TTC workout had a time limit to complete all work), all workout scores were converted into a repetition completion rate (i.e., repetitions completed divided by TTC or workout duration; repetitions□minute^-1^), as previously described (29). Regardless of the specific workout’s scoring format, a greater repetition completion rate would always indicate a better competition score. This metric was also used to calculate each athlete’s percent rank within each workout.

### Statistical Analysis

Initially, the assumption of normal distribution was verified by the Shapiro-Wilk test. Subsequently, separate two-tailed, two-way (Time x Sex) analyses of variance with repeated measures were performed for each repeated CFO workout comparison. Dependent variables included age and original overall rank (within each year) and original workout rank, calculated percent rank, and repetition completion rate. Except for the instance when a CFO workout was repeated twice (i.e., 11.6 vs. 12.5 vs. 18.5), sphericity was assumed for all repeated workout comparisons. For the exception, the Greenhouse-Geisser correction was applied because sphericity was violated on each comparison. All significant main effects and interactions were further assessed by pairwise comparisons using the Bonferroni adjustment. Effect sizes (η^2^_P_: Partial eta squared) were also used to quantify the magnitude of any observed differences (30). Interpretations of effect size were evaluated at the following levels: small effect (0.01-0.058), medium effect (0.059-0.137) and large effect (> 0.138). All statistical analyses were performed using SPSS (v. 28.0, SPSS Inc., Chicago, IL). Significance was accepted at an alpha level of *p* ≤ 0.05, and all data are reported as mean ± SD.

## RESULTS

### Ranking and athlete age

No differences were seen in the workout percent rank calculated for each repeated workout within each comparison, except for a main effect for sex when comparing 12.4 and 13.3 (F = 17, *p* < 0.001, η^2^_P_= 0.02)., Percent workout rank was greater in men (29.8 ± 27.1%) than women (27.1 ± 23.5%) during the workout’s original and repeated appearances. In contrast, significant time x sex interactions (F = 14 – 1089, *p* < 0.001, η^2^_P_= 0.01 – 0.52) were noted with original overall rank for each workout comparison. In each comparison, men ranked lower within their division than women, and ranking further declined whenever a CFO workout was repeated. Likewise, the original rank assigned for performance in each workout followed a similar pattern in each comparison.

Significant time x sex interactions were only found in the comparisons between 11.1 and 14.1 (F = 4, *p* = 0.034, η^2^_P_< 0.01) and between 11.6, 12.5, and 18.5 (F = 3, *p* = 0.041, η^2^ _P_< 0.01). Average age in men increased from 2011 to 2012 (+1.1 years, *p* = 0.015), from 2011 to 2014 (+ 2.0 years, *p* < 0.001), and from 2012 to 2018 (+1.8 years, *p* < 0.001), but only from 2011 to 2018 in women (+1.5 years, *p* < 0.001). Main effects for time, where age increased in subsequent years, were noted in all remaining comparisons except for between 14.5 and 16.5 (*p* = 0.053). Main effects for sex were also noted with the comparisons between 14.2 and 15.2 (F = 9, *p* = 0.003, η^2^_p_ = < 0.01), 14.5 and 16.5 (F = 5, *p* = 0.023, η^2^_p_ < 0.01), and 16.2 and 19.2 (F = 6, *p* = 0.013, η^2^_p_ < 0.01), where men were older than women in each case. Comparisons between sexes across repeated CFO workouts for age, overall rank, workout rank, and workout percentile rank are presented in Table 4.

**Table 4.**
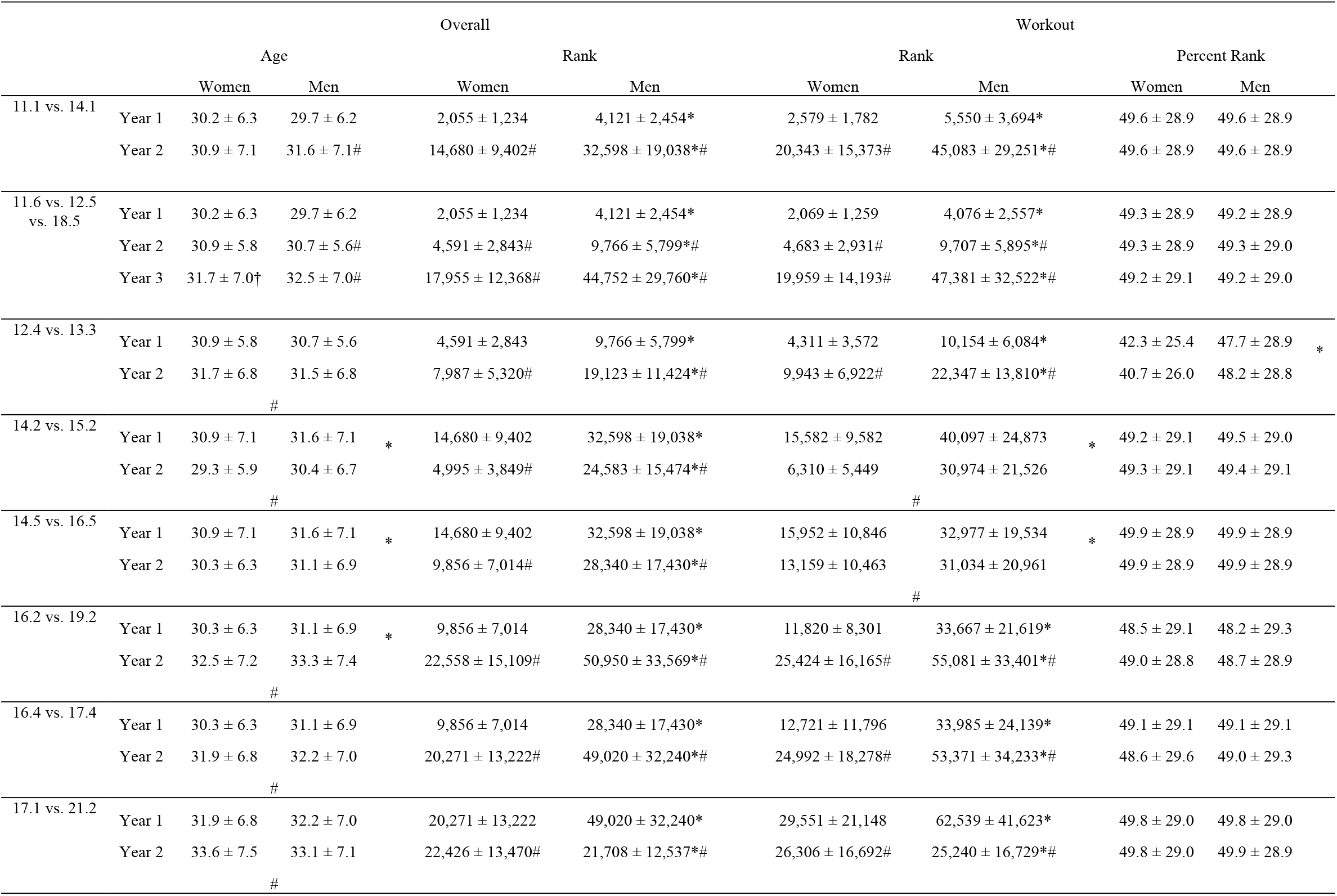
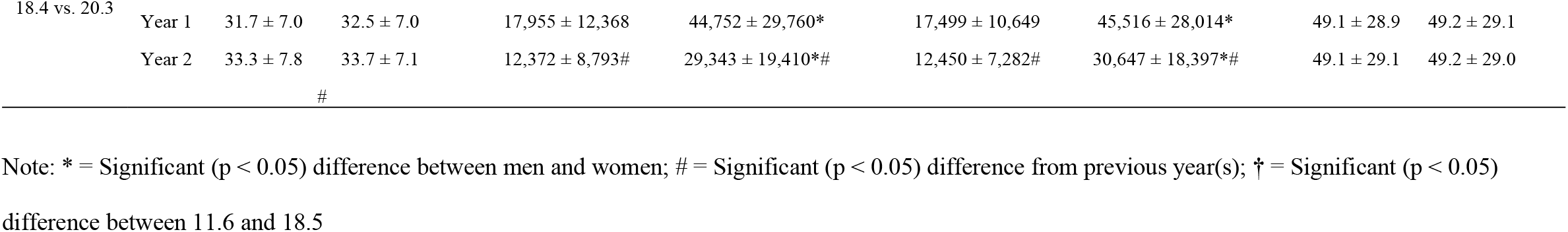
Comparisons between age, overall rank, workout rank, and workout percent rank in repeated CFO workouts.

### Workout completion rate

Significant time x sex interactions (F = 14 – 357, *p* < 0.001, η^2^ _P_= 0.01 – 0.26) were observed for all workout completion rate comparisons except for between 18.4 and 20.3, where no differences were found between sexes or repeated performance. Comparisons between sexes and across repeated CFO workouts for repetition completion rate are illustrated in Figure 1.

**Figure 1.**
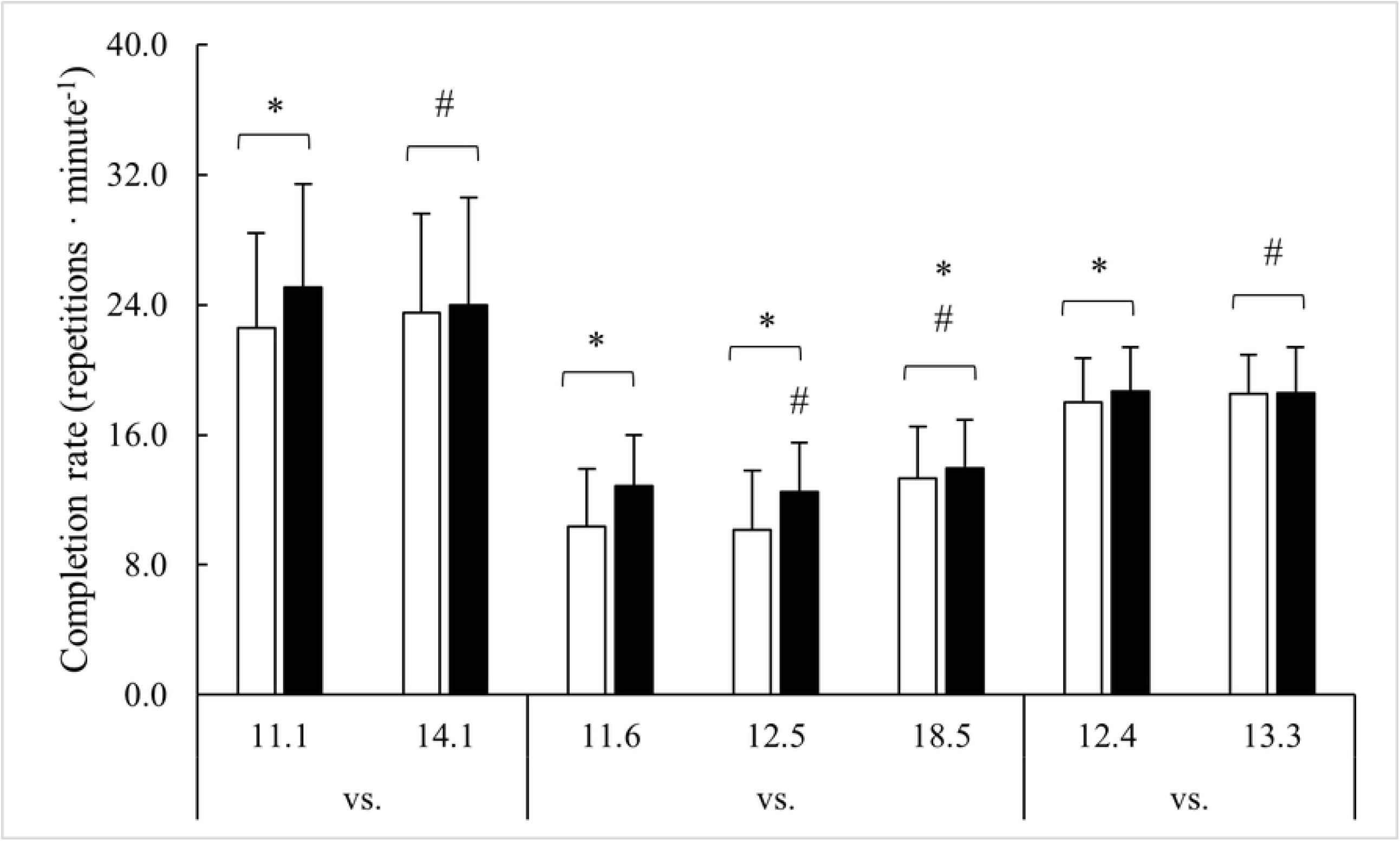
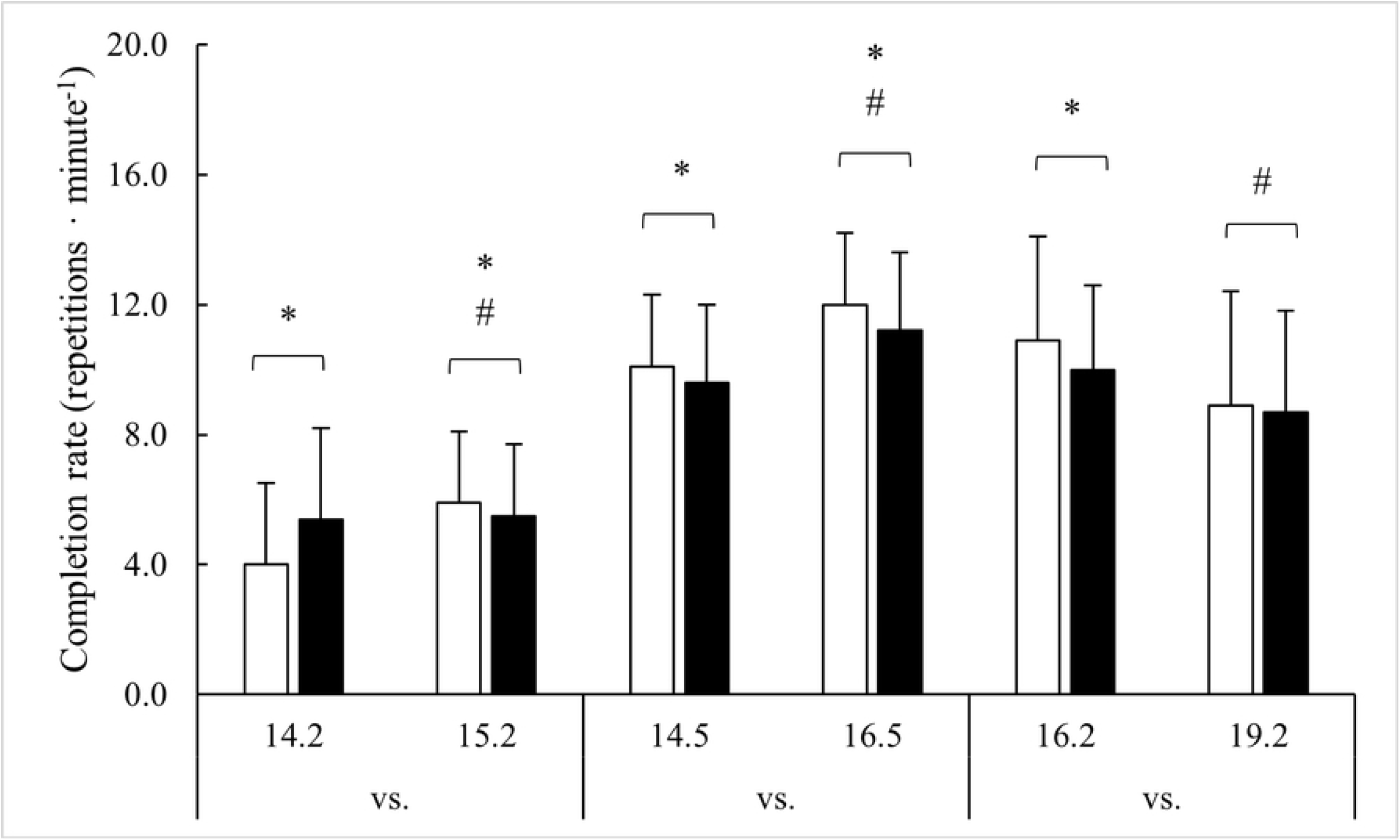
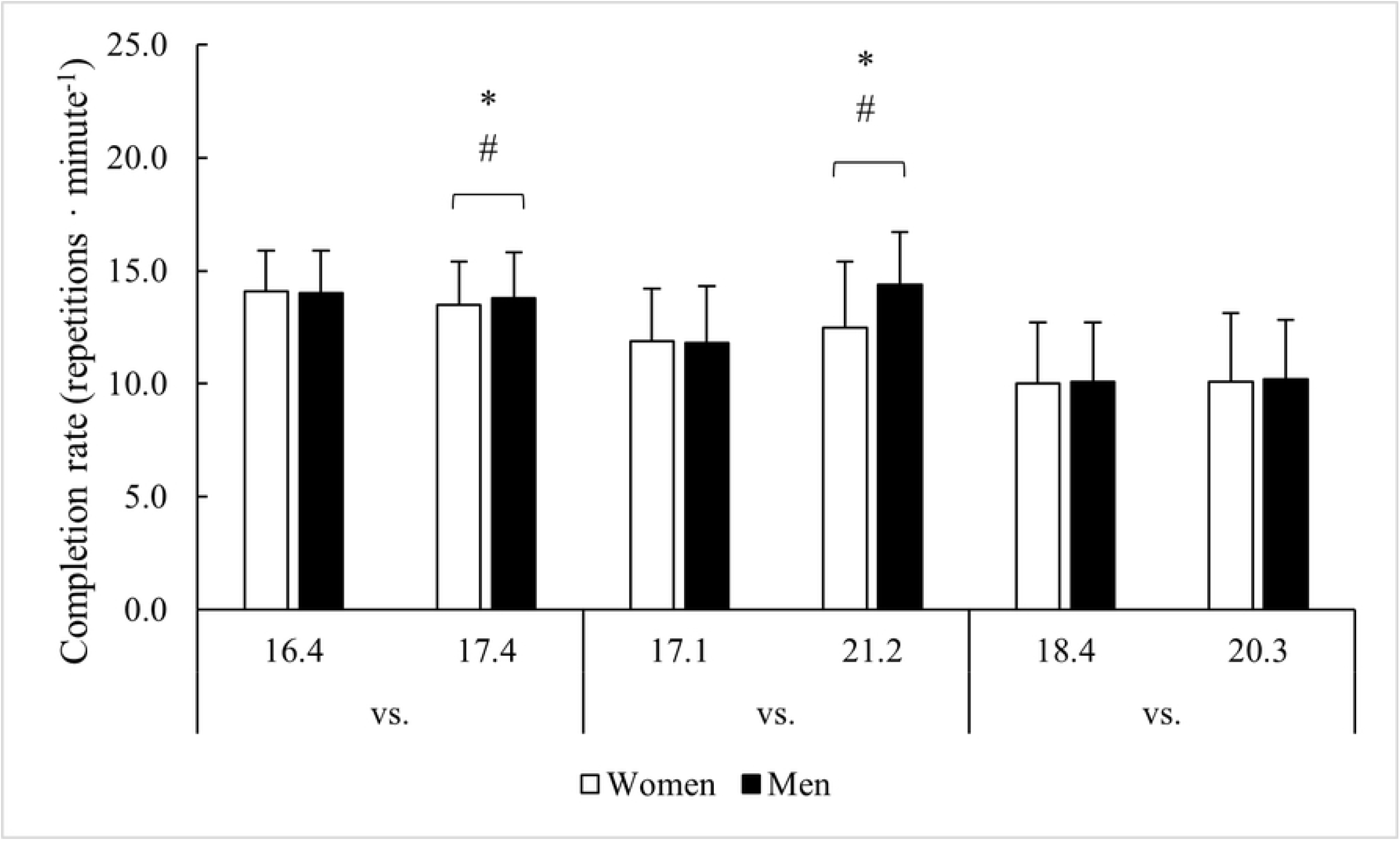
Repeated workout comparisons for repetition completion rate. Note: * = Significant (*p* < 0.05) difference between men and women; # = Significant (*p* < 0.05) difference from previous year(s).

#### 11.1 vs. 14.1

Though men completed 11.1 at a faster rate than women (+2.5 repetitions □ minute^-1^, *p* < 0.001), no differences were seen in 14.1. In 2014, women improved repetition completion rate (+4.0%, *p* < 0.001), while men declined (–4.4%, *p* < 0.001).

#### 11.6 vs. 12.5 vs. 18.5

Men completed the workout faster than women in each year (0.6 – 2.5 repetitions □ minute^-1^, *p* < 0.001). However, compared to 2011, their performance declined in 2012 (–3.0%, *p* = 0.002) before improving by 8.4% in 2018 (*p* < 0.001). Women remained steady from 2011 to 2012 before improving in 2018 (+28.4%, *p* < 0.001).

#### 12.4 vs. 13.3

Men completed 12.4 at a faster rate than women (0.7 repetitions □ minute^-1^, *p* < 0.001) but not 13.3 (*p* = 0.703). In 2013, women improved their repetition completion rate by 0.5 repetitions □ minute^-1^, *p* < 0.001) and men slowed by 0.1 repetitions □ minute^-1^ (*p* < 0.001).

#### 14.2 vs. 15.2

Men completed 14.2 at a faster rate than women (1.4 repetitions □ minute^-1^, *p* < 0.001) but slowed for 15.2 by 0.2 repetitions □ minute^-1^ (*p* < 0.024). Women improved their repetition completion rate for 15.2 by 1.9 repetitions □ minute^-1^ (*p* < 0.001) and exceeded the pace in men by 0.4 repetitions □ minute^-1^ (*p* = 0.004).

#### 14.5 vs. 16.5

Compared to men, a faster repetition completion rate was seen in women for both 14.5 (0.5 repetitions □ minute^-1^, *p* < 0.001) and 16.5 (0.8 repetitions □ minute^-1^, *p* < 0.001), with both men (1.6 repetitions □ minute^-1^, *p* < 0.001) and women (1.9 repetitions □ minute^-1^, *p* < 0.001) improving their speed from 2014 to 2016.

#### 16.2 vs. 19.2

A faster repetition completion rate was seen in women for 16.2 (0.9 repetitions □ minute^-1^, *p* < 0.001) but not 19.2 (*p* = 0.360). Though men performed 19.2 at a slower pace (–1.2 repetitions □ minute^-1^, *p* < 0.001), women experienced a greater decline (–2.0 repetitions □ minute^-1^, *p* < 0.001).

#### 16.4 vs. 17.4

No differences in repetition completion rate were noted between men and women for 16.4 (*p* = 0.680). In 2017, men completed 17.4 at a faster rate than women (0.2 repetitions □ minute^-1^, *p* < 0.047), but this was due to men slowing down less (–0.3 repetitions □ minute^-1^, *p* < 0.001) than women (–0.6 repetitions □ minute^-1^, *p* < 0.001).

#### 17.1 vs. 21.2

No differences in repetition completion rate were noted between men and women for 17.1 (*p* = 0.441). In 2021, men completed 21.2 at a faster rate than women (2.0 repetitions □ minute^-1^, *p* < 0.047), and this was due to men improving their pace by 2.6 repetitions □ minute^-1^ (*p* < 0.001) compared to the 0.5 repetitions □ minute^-1^ improvement (*p* < 0.001) seen in women.

## DISCUSSION

This study examined performance changes in repeated CFO workouts and investigated whether men and women performed differently across time. A secondary aim was to determine how changes in performance affected competition ranking. The data indicated that out of nine separate repeated CFO workouts, repetition completion rate improved in five, slowed in two, and in one, remained the same (i.e., the 9-minute AMRAP programmed for 18.4 and 20.3). Though pace generally improved over time, the overall rank awarded declined. An outcome most likely due to increased participation over time because neither overall nor workout percent rank changed. Nevertheless, to achieve a comparable or better rank in a repeated CFO workout, athletes had to improve performance beyond what was typically seen across all competitors.

Additionally, initial, and subsequent performances were not equal among men and women. Men were initially faster in four of the workouts, while women outpaced men in two workouts, and tied in the remaining three. Men still demonstrated a faster pace in more workouts when they were repeated (3 workouts versus 2 workouts), but the specific combinations of workouts where they were faster differed slightly. Men and women tied on the second iteration of two workouts where men were initially faster and were slower than women in a third workout where they had initially been faster. Women also tied men on the second iteration of a workout where they were initially faster.. Aside from a preliminary conference presentation (27), this is the first study to investigate performance changes in official CrossFit® competition workouts.

Improved performance was observed in six of the nine repeated CFO workouts. A learning effect would seem to be the most likely explanation for these improvements. During the CFO, athletes are given a 4-day window to submit their best score after each workout’s release (5). A better score is accomplished by employing the fastest sustainable (for the duration of the workout) pace that simultaneously manages fatigue and maximizes workout density (i.e., repetitions □ minute^-1^) (29). Since each CFO workout is novel (except for those examined in this study) (4, 5), finding one’s optimal pace within four days may be difficult. Ideally, athletes would reconcile their physiological and skill-related abilities with a workout’s exercise complexity, relative intensity, workload requirements, and structural design (i.e., AMRAP, TTC) on their first attempt. Though experience may facilitate this process (7, 12-14), each workout presents a unique set of conditions that might necessitate multiple attempts to find an optimal pace. Each additional workout (or attempt) would contribute to accumulated fatigue and damage that might not diminish within the submission window (31), and this could negatively impact effort on subsequent attempts. Therefore, it is possible for athletes to not find their ideal pacing strategy for each workout within a single CFO competition. However, after their introduction, CFO workouts may be integrated into training and/or frequently discussed (in-person or online) amongst athletes. With an average of 2.4 years separating repeated CFO workouts (18, 19), athletes have ample opportunities to refine their pacing strategy. Albeit they would not know which and when specific CFO workouts might be repeated, leaving an ever-growing list of potential workouts in need of practice.

Having time to perfect pacing strategy may have been less relevant to performance in the three workouts that did not improve (i.e., 16.2 – 19.2, 16.4 – 17.4, and 18.4 – 20.3). Compared to all other workouts, these three required more strength and capacity to lift heavier loads or perform complex gymnastic movements for multiple repetitions. For instance, deadlift loads ranged from 225 – 315 lbs. (102.1 – 142.9 kg) for men and 155 – 205 lbs. (70.3 – 93.0 kg) for women, and power clean loads ranged from 135 – 315 lbs. (61.2 – 142.9 kg) for men and 85 – 205 lbs. (38.6 – 93.0 kg) for women. Further, these heavier loads were always paired with one or more gymnastics-calisthenic movements prescribed for multiple repetitions (e.g., 25 toes-to-bar, 50 double-unders, 45 – 55 handstand push-ups, and 150 feet [45.7 m] of handstand walking). In contrast, the highest load required in all other workouts was for thrusters (men: 100 lbs. [45.4 kg]; women: 65 lbs. [29.5 kg]), and these were typically paired with low-complexity calisthenics (e.g., burpees, burpee box jumps, and double-unders) prescribed at noticeably less volume. The only instance where gymnastics prescription was comparable involved 30 muscle ups (i.e., 12.4 – 13.3), and the average athlete did not even complete a full round of this workout. While modifying pacing would seem to have a more immediate effect on these latter workouts, improving upon the former would have required more time to develop strength and strength endurance, as well as acquire or improve upon relevant gymnastic skills (32-34). On average, athletes had less time to work on relevant skills for these three repeated workouts (2.0 years) compared to the other workouts (2.6 years).

An alternate hypothesis for explaining the observed changes in performance might involve the relative growth of the sport. Compared to the first CFO in 2011, the number of athletes meeting this study’s criteria increased 156 – 1305% and 168 – 1138% for women and men, respectively. Men peaked in 2019, while participation in women peaked in 2021. Similar trends were noted for all participating athletes (i.e., including scaled and age-group divisions). The competitive aspect about CrossFit® has been identified as a highly influential factor for participation and retention (35). However, little is known about the athletic and physical activity backgrounds among pre-existing and newly-introduced CFO participants. In a 2016 epidemiological survey of Brazilian CrossFit® athletes (36), only 6.2% of respondents stated they had no sports experience prior to starting CrossFit®. Rather, 70.5% reported being physically active on > 3 days per week doing a variety of activities (i.e., weight training [72.1%], running [36.9%], soccer [19.2%], and martial arts [18.3%]), and most (67.0%) had been doing this for more than 3 years. Skills learned and developed across various sports and levels of competition are well known to have value to an athlete’s primary sport (16, 17). It is possible that improved CFO performance may at least be partially due to newcomers being drawn from other sports and already possessing aptitude in many of the skills and traits needed for success.

An intentional examination into the development of relevant skills, and whether they were obtained prior to or during CrossFit® training, would provide greater insight into the factors responsible for the growth of this sport.

The factors responsible for the growth in CFO participation may also be relevant to the differences seen between men and women. In 2011, women accounted for 34.3% of all competitors (or 30.2% of competitors meeting this study’s criteria). Out of all the instances where men outpaced women, half (*n* = 4) were seen during initial or repeated CFO workouts appearing within the competition’s first two years (i.e., 11.1, 11.6, 12.4, and 12.5). Since 2011, the percentage of competitors who were women increased by 0.8% per year; a 0.3% increase per year out of competitors who met this study’s criteria. Concomitantly, an equal number of instances (initial and repeated) where a performance advantage was observed either for men (*n* = 4) or women (*n* = 4) were seen during this time. No advantages were observed in all other instances (*n* = 7). Moreover, out of the four workouts where men initially held an advantage, women either eliminated (12.4 – 13.3 and 11.1 – 14.1) or claimed the advantage (14.2 to 15.2). Thus, female CFO participation is clearly increasing, and women appear to be experiencing greater improvements. Indeed, average performance improvement across all workouts was 8.3% for women compared to 2.8% in men. The driving force(s) behind these improvements is/are not well understood. It is possible that this observation is a simple mathematical function where lower values do not require dramatic additions in absolute numbers to experience larger percent increases. The potential lure of CrossFit® and competition may have also drawn women with athletic backgrounds to the overall athlete pool. That said, men appear to place more importance on the competitive aspects of CrossFit® (37) whereas women seem to more commonly be drawn by the social and health (physical and mental) related benefits (37, 38). Instead, it may be speculated that increased female participation and performance are more closely related to a growing realization that this sport empowers women unlike more traditional sports and physical activity settings (39, 40). That in many cases, women can perform as good or better than men.

The presence and adequacy of program scaling are the caveats to the observed sex differences on pacing. There appears to be an emphasis placed on scaling workout characteristics (4, 5), and this is presumably meant to acknowledge known physiological differences and elicit a similar challenge to both men and women (4, 5, 21). If applied correctly, significant differences in repetition completion rate should not exist. However, performance differences were noted, as were inconsistencies with scaling application across exercise types. For example, resistance training loads, wall ball shot medicine ball weight and target distance, and box jump height were all scaled, but not gymnastic and calisthenic movements (e.g., chest-to-bar pull-ups, handstand push-ups, handstand walking, burpees, rope jumping), continuous exercise patterns (referred to as monostructural; e.g., rowing), or programming durations. Although scaled loads might account for strength differences (22), their arbitrary prescription (i.e., loads assigned to women were 67.1% [range = 62.2 – 73.3%] less than those assigned to men) assumes a specific strength difference that cannot be known across thousands of competitors. Likewise, it seems that known differences in body mass (23) provide the basis for why gymnastics and calisthenics are not scaled, but this rationale fails to account for known upper-body strength differences (22) relevant to several exercises. Finally, the potential sex differences with aerobic and anaerobic capacity (22, 24) are not addressed by scaling continuous exercises or workout durations. Thus, it can be concluded that scaling was not adequately applied across all CFO workouts. That said, the present study limited its focus to repeated CFO workouts, not all CFO or competition workouts. It was not adequately designed to make definitive conclusions scaling adequacy. Nevertheless, our findings on sex differences in programming and performance warrant further investigation.

The present study’s findings are not without limitations. Competition rule and structure changes (e.g., specific days allotted to submit scores, number of workouts, submission card details), as well as variations in when each workout iteration appeared (e.g., 18.4 appeared on week 4 of 2018, whereas 20.3 occurred on week 3 of 2020), and the number of attempts individual athletes made before submitting their best score, have all affected the exact context though which each workout was performed. Nevertheless, these are unavoidable variations that are consistent with the training strategy (2) but should still be kept in mind when interpreting these results. Another limitation involved purposeful, stratified selection of cases based on percent rank to make fair comparisons between sexes and repeated performances. The non-random stratification was done to best approximate a normally-distributed population (28) and since repeated performances were not followed in a random sample of the same exact athletes, it is possible that individual differences and variances in age and relevant physical, physiological, and psychological traits influenced the analyzed scores (6-14). However, given the growth in CFO participation, it would not have been possible to follow a sufficient sample of the same athletes over a decade of competitions. Even if it were, that sample might be more accurately representative of a specific subset of more experienced CFO athletes, rather than a more heterogenous representation of typical CFO athlete experience. Conversely, the opposite rationale underpinned this study’s exclusion criteria. Cases were excluded if the athlete did not earn a score beyond a minimum threshold assigned to each CFO workout of a specific year.

These measures were meant to ensure that our findings were representative of a homogenous sample of healthy (i.e., those that did not miss a workout due to injury), well-rounded (i.e., non-workout specialists or those attempting to boost their rank by only completing a small number of repetitions in a specific workout) CrossFit® athletes. But in doing so, it is possible that representative, low-ranking cases were eliminated, and this could have slightly skewed our results. Finally, the validity of the extracted scores is ultimately reliant on determinations made by CFO competition officials (5). As these cannot be verified, it is possible that the analyzed scores included errors in reporting, individual variation in meeting exercise movement standards, and outright cheating. Still, the conservative approach in estimating expected sample requirements for sufficient statistical power, and then exceeding the minimum sample size should have limited the impact of these limitations on our results.

The findings of this study suggest that performance (measured as repetition completion rate) in most repeated CFO workouts, particularly females, has improved since the competition’s inception. These data serve as initial documentation that, though often scaled, men and women scored differently in ∼63.2% of workouts. These differences warrant a more in-depth look across a broader range of workouts, and more specifically, how they might affect acute and long-term physiological responses. Doing so, might help guide more effective, sex-equated scaling practices. From a competitive standpoint, the general improvements seen among athletes, along with increased participation, have made it more difficult for athletes to improve their overall rank. Athletes might maintain their percent rank but drop in overall ranking even if they complete repeated workouts at a faster rate. In this regard, maintaining one’s overall rank across iterations of a workout should be viewed as a positive outcome, as it would imply greater improvements compared to the average competitor. Ranking higher appears to require the individual athlete to improve their pacing to an even greater degree than the remainder of the competitive field. Athletes and coaches are advised to maintain perspective when identifying areas in need of attention, and to focus on finding suitable pacing strategies that balance efficiency with physiological attributes when attempting to improve performance in specific workouts.

## Data Availability

All data is publicly available at: https://games.crossfit.com/leaderboard/open/2023?view=0&division=2&region=0&scaled=0&sort=0

## REFERENCES

1. Feito Y, Heinrich K, Butcher S, Poston W. High-Intensity Functional Training (HIFT): Definition and Research Implications for Improved Fitness. Sports. 2018;6(3):76.

2. Glassman G. CrossFit training guide level 1.: The CrossFit Journal; 2011.

3. CrossFit. Finding the Fittest on Earth. CrossFit Games [Internet]. 2022 September 15, 2022. Available from: https://games.crossfit.com/history-of-the-games.

4. CrossFit. Open Workouts. CrossFit Games [Internet]. 2021; (August 31). Available from: https://games.crossfit.com/workouts/open/2021.

5. CrossFit. Games Competition Rulebook: The CrossFit Journal; 2022.

6. Butcher SJ, Neyedly TJ, Horvey KJ, Benko CR. Do physiological measures predict selected CrossFit® benchmark performance? Open Access Journal of Sports Medicine. 2015;6:241.

7. Bellar D, Hatchett A, Judge L, Breaux M, Marcus L. The relationship of aerobic capacity, anaerobic peak power and experience to performance in CrossFit exercise. Biol Sport. 2015;32(4):315–20.

8. Feito Y, Giardina MJ, Butcher S, Mangine GT. Repeated anaerobic tests predict performance among a group of advanced CrossFit-trained athletes. Applied Physiology, Nutrition, and Metabolism. 2018;44(7):727–35.

9. Dexheimer JD, Schroeder ET, Sawyer BJ, Pettitt RW, Aguinaldo AL, Torrence WA. Physiological Performance Measures as Indicators of CrossFit® Performance. Sports. 2019;7(4):93.

10. Zeitz EK, Cook LF, Dexheimer JD, Lemez S, Leyva WD, Terbio IY, et al. The relationship between Crossfit® performance and laboratory-based measurements of fitness. Sports. 2020;8(8):112.

11. Carreker JDD, Grosicki GJ. Physiological predictors of performance on the CrossFit® “Murph” challenge. Sports. 2020;8(7):92.

12. Mangine GT, Tankersley JE, McDougle JM, Velazquez N, Roberts MD, Esmat TA, et al. Predictors of CrossFit Open performance. Sports. 2020;8(7):102.

13. Mangine GT, McDougle JM. CrossFit® open performance is affected by the nature of past competition experiences. BMC Sports Science, Medicine and Rehabilitation. 2022;14(1):1–15.

14. Mangine GT, Mcdougle JM, Feito Y. Relationships Between Body Composition and” Fran” Performance are Modulated by Competition Class and Skill. Front Physiol. 2022:969.

15. Krakauer JW, Hadjiosif AM, Xu J, Wong AL, Haith AM. Motor Learning. Comprehensive Physiology. 2019;9(2):613–63.

16. Brenner JS. Sports specialization and intensive training in young athletes. Pediatrics. 2016;138(3).

17. Myer GD, Jayanthi N, DiFiori JP, Faigenbaum AD, Kiefer AW, Logerstedt D, et al. Sports specialization, part II: alternative solutions to early sport specialization in youth athletes. Sports Health. 2016;8(1):65–73.

18. Micklewright D, Parry D, Robinson T, Deacon G, Renfree A, St Clair Gibson A, et al. Risk perception influences athletic pacing strategy. Med Sci Sports Exerc. 2015;47(5):1026–37.

19. Santalla A, Naranjo J, Terrados N. Muscle efficiency improves over time in world-class cyclists. Med Sci Sports Exerc. 2009;41(5):1096–101.

20. Leaderboard. Leaderboard 2021 [Available from: http://games.crossfit.com/leaderboard.

21. Laxton K. Closing the gender gap - Empowering women in sport. CrossFit Journal [Internet]. 2022 September 15, 2022. Available from: https://games.crossfit.com/article/closing-gender-gap-how-crossfit-empowers-women-spor.

22. Sandbakk ø, Solli GS, Holmberg H-C. Sex differences in world-record performance: the influence of sport discipline and competition duration. Int J Sports Physiol Perform. 2018;13(1):2–8.

23. Huebner M, Perperoglou A. Sex differences and impact of body mass on performance from childhood to senior athletes in Olympic weightlifting. PLoS One. 2020;15(9):e0238369.

24. Hunter SK. The relevance of sex differences in performance fatigability. Med Sci Sports Exerc. 2016;48(11):2247.

25. Toledo R, Dias MR, Toledo R, Erotides R, Pinto DS, Reis VM, et al. Comparison of Physiological Responses and Training Load between Different CrossFit® Workouts with Equalized Volume in Men and Women. Life. 2021;11(6):586.

26. Mangine GT, Grundlingh N, Feito Y. Normative Scores for CrossFit® Open Workouts: 2011–2022. Sports. 2023;11(2):24.

27. Mangine GT, editor Sex differences and performance changes over time in a repeated fitness competition workout containing thrusters and chest-to-bar pull-ups. National Strength & Conditioning Association National Conference; 2022; New Orleans, LA.

28. Weir JP, Vincent WJ. The Normal Curve. Statistics in Kinesiology. 5th ed. Champaign, IL: Human Kinetics; 2021. p. 55–65.

29. Mangine GT, Feito Y, Tankersley JE, McDougle JM, Kliszczewicz BM. Workout Pacing Predictors of Crossfit Open Performance: A Pilot Study. Journal of Human Kinetics. 2021;78(1):89–100.

30. Cohen J. Statistical Power Analysis for the Behavioral Sciences. Routledge. 1988:284-8.

31. Tibana RA, Prestes J, Nmf DES, Vc DES, o DETN, Baffi M, et al. Time-Course of Changes in Physiological, Psychological, and Performance Markers following a Functional-Fitness Competition. International Journal of Exercise Science. 2019;12(3):904–18.

32. Gulbin J, Weissensteiner J, Oldenziel K, Gagné F. Patterns of performance development in elite athletes. European journal of sport science. 2013;13(6):605–14.

33. Ikezoe T, Kobayashi T, Nakamura M, Ichihashi N. Effects of Low-Load, Higher-Repetition vs. High-Load, Lower-Repetition Resistance Training Not Performed to Failure on Muscle Strength, Mass, and Echo Intensity in Healthy Young Men: A Time-Course Study. The Journal of Strength & Conditioning Research. 2020;34(12):3439–45.

34. Shemmell J, Tresilian JR, Riek S, Carson RG. Musculoskeletal constraints on the acquisition of motor skills. Skill Acquisition in Sport: Routledge; 2004. p. 414–32.

35. Fisher J, Sales A, Carlson L, Steele J. A comparison of the motivational factors between CrossFit participants and other resistance exercise modalities: a pilot study. J Sports Med Phys Fitness. 2017;57(9):1227–34.

36. Sprey JW, Ferreira T, de Lima MV, Duarte Jr A, Jorge PB, Santili C. An epidemiological profile of CrossFit athletes in Brazil. Orthopaedic journal of sports medicine. 2016;4(8):2325967116663706.

37. Bycura D, Feito Y, Prather C. Motivational factors in CrossFit® training participation. Health Behavior and Policy Review. 2017;4(6):539–50.

38. Coyne P, Woodruff SJ. Examining the influence of CrossFit participation on body image, self-esteem, and eating behaviours among women. Journal of Physical Education and Sport. 2020;20(3):1314–25.

39. Schrijnder S, van Amsterdam N, McLachlan F. ‘These chicks go just as hard as us!’(Un) doing gender in a Dutch CrossFit gym. International Review for the Sociology of Sport. 2021;56(3):382–98.

40. Washington MS, Economides M. Strong is the new sexy: Women, CrossFit, and the postfeminist ideal. Journal of Sport and Social Issues. 2016;40(2):143–61.

